# Participatory prototyping of a tailored U=U (undetectable=untransmittable) message to increase HIV testing in men in Western Cape, South Africa

**DOI:** 10.1101/2021.05.03.21256402

**Authors:** Philip Smith, Dvora L. Joseph Davey, Laura Schmucker, Cal Bruns, Linda-Gail Bekker, Andrew Medina-Marino, Harsha Thirumurthy, Alison Buttenheim

## Abstract

**Introduction:** Taking daily ART eliminates sufficient virus so that HIV is undetectable via viral load (VL) testing within 24 weeks. HIV-positive individuals with an undetectable VL cannot transmit HIV to sexual partners or through giving birth, a message commonly referred to as U=U (undetectable equals untransmittable). Since South African men have poorer HIV outcomes than women, we used interactive human centred design co-creation workshops to ask men from high HIV burden communities in Cape Town, South Africa to create a U=U message aimed at increasing HIV testing and ART uptake in men.

**Methods:** Two facilitators explained the U=U message to the men (n =39) attending the workshop and asked them how to effectively communicate the message. Participants designed messages to assuage fears of testing HIV positive, explaining that ART enables HIV positive people to live normally and makes the virus “untransmittable” to their sexual partners.

**Results:** Participants developed three insights for the U=U message; 1) “Introduce” the modern antiretroviral pill, 2) positively redefine the man for whom the pill is intended, and 3) simplify the benefits of ART for men. Participants’ messages emphasised 1) “*you cannot spread the virus (HIV) to the other person*” 2) and “*(the pill) keeps on killing the virus so I can live a normal life for the rest of my life*.”

**Discussion:** Men in the workshops co-created a simple U=U message to address fears of testing HIV positive, emphasising the pill’s positive effects. Co-created, tailored messaging may improve the uptake of HIV services for South African men.

## Introduction

South African males, compared with their female counterparts, have lower rates of HIV testing, ART start (antiretroviral treatment initiation), and mortality on treatment (1–5). This disparity results in more costly interventions to manage opportunistic comorbidities (6,7). High disease burden communities require interventions that will increase testing in men who are at risk of HIV acquisition and onward transmission to achieve viral suppression both individually and within high-risk communities (8,9).

Taking daily ART eliminates sufficient virus so that HIV is undetectable via viral load (VL) testing within 24 weeks (10). HIV-positive individuals with an undetectable VL cannot transmit HIV to sexual partners or through giving birth, a message commonly referred to as U=U (undetectable equals untransmittable). U=U messaging has been implemented worldwide, with studies showing the acceptability and impact of the message on HIV testing, ART start, and VL suppression (11). While studies have investigated acceptability among key populations, one study conducted in 25 countries found that awareness was highest among men who have sex with men, followed by women who have sex with men (11). In one qualitative study conducted with women in South Africa, participants suggested that emphasising the importance of taking care of oneself was a valued strategy for improving treatment adherence (12). A study conducted with men from rural areas in South Africa found that while the men were aware of the positive effects of ART on “regaining one’s health”, improving appearance, and increasing longevity, few of the participants were aware of the preventive effects of HIV treatment (13). Moreover, once the participants were made aware of the effects on transmission, they offered that knowing this would motivate men to test for HIV and could improve adherence.

While the message has been recommended for the endemic South African setting, there is, however, limited evidence on the effect of the message in the South African population. Additionally, since men have poorer uptake of HIV services and poorer health and morbidity outcomes compared women, it may be ideal to develop and test the impact of a U=U message that has been tailored for the context. The aim of this project was to recruit South African men living in a high HIV burden community in Cape Town to co-create a U=U message with the purpose of increasing HIV testing and ART start or re-engagement among their peers.

## Methods

### Setting and participants

The message development was conducted with men 18-49 years old from the Klipfontein Mitchells Plain (KMP) District in Cape Town, South Africa. The district has a high HIV disease burden and has high density population. A trained recruiter invited men from high traffic locations to participate in an interactive human-centered design (HCD) co-creation workshop about HIV treatment, HIV testing and barriers to both. HCD has been used to co-create tailored products and services with end users for whom they are intended (14). This method has been used for HIV testing, prevention, and treatment services (15). The trained facilitator presented the theory behind the U=U message with the goal of developing a U=U message for their peers. The facilitators presented the U=U message to the participants, stating that people living with HIV who take ART daily will reduce the virus to the extent that they will not infect their sexual partners. Participants were reimbursed (in South African RAND) for their participation in the workshop.

### Design

The workshops were designed to develop a U=U message using a participatory, human-centred design framework. There is increasing recognition that intervention design should incorporate viewpoints from the target audience to enhance collaboration, improve acceptability, and increase uptake (16,17). This participatory research included two workshops from a population of men that may potentially be recipients of the intervention.

### Process

The study team recruited men from high foot traffic locations in the KMP community to participate in HCD workshops for the development of a locally relevant and resonant U=U message that even if diagnosed as living with HIV, they can be untransmittable if they take treatment daily, and no longer transmit the virus to sex partners. The goal was to detraumatize HIV infection with a clear picture of a normal future that could mitigate the fears of testing HIV positive. Each workshop participant was asked to give informed consent, and then a trained facilitator presented in layman’s terms the scientific rationale and evidence substantiating the U=U message (Figure 1). The facilitators then stated that the intention was to contextualise the message for the local context and elicited insights from the men on how to do this. Specifically, the facilitators asked 1) what might I hear to make me more curious about this pill, 2), what might we do to convince everyone in my suburb that I’m HIV safe thanks to this pill, 3) what might we say to make you confident that I’m HIV safe thanks to this pill, and 4) how might your favourite brand sell this pill to men like you? These questions were displayed on posters in the workshop venue (Figure 2). Participants were then facilitated through a series of HCD group exercises designed to co-create a face-to-face presentation of this information (a 1 minute “sales pitch”) that these men believed would resonate with their community peers in language, tone and structure.

**Figure 1:**
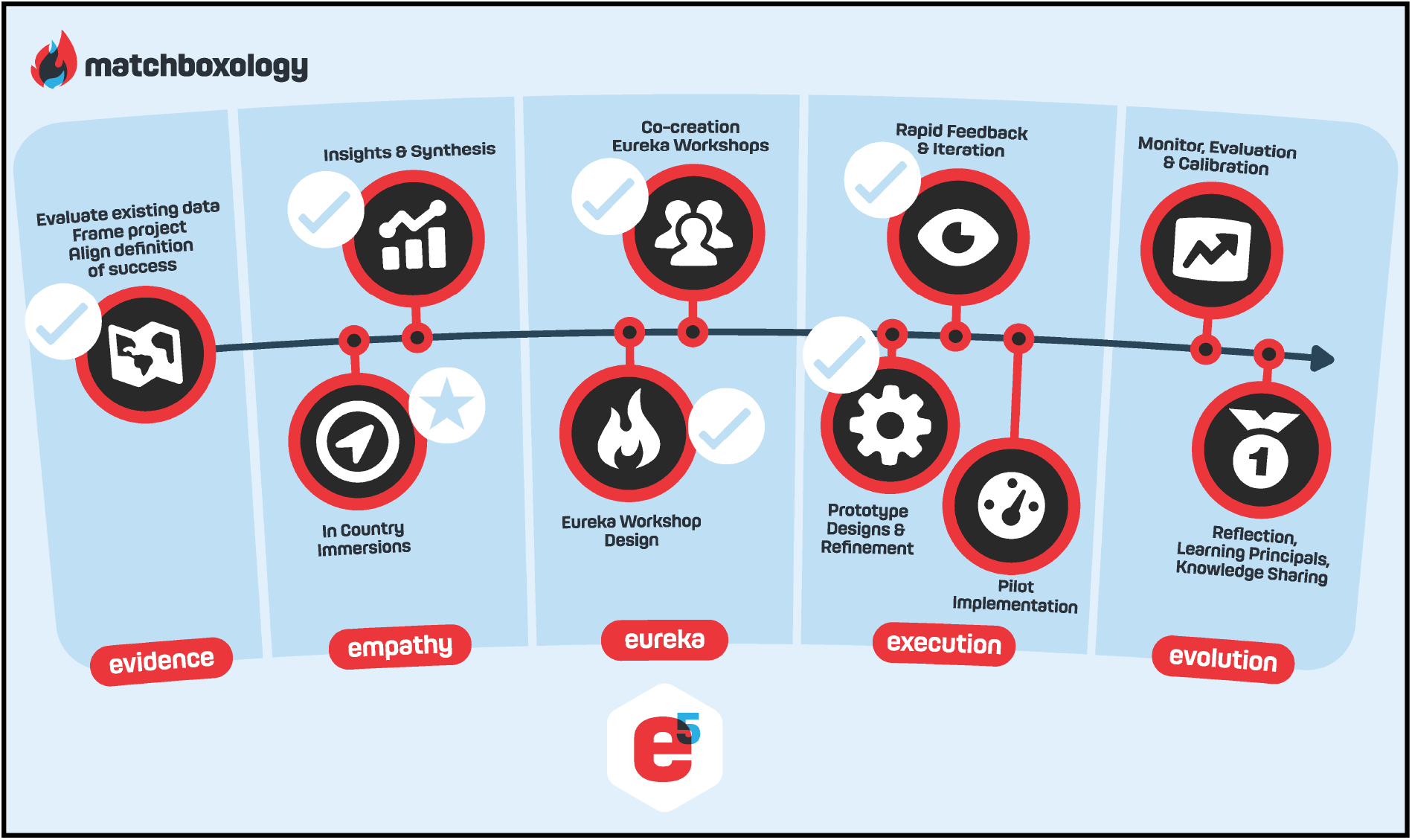
human-centred design workshop process.

**Figure 2.** **Visual prompts for workshops (top). Images from workshops (bottom) (please request images from author)**

The men presented their initial suggestions as “sales pitches”. Participants self-critiqued their first try and based on suggestions, refined their pitch, and again presented this again, at which time this was audio recorded and transcribed (Figure 1).

These final pitches were ranked by participants in order of perceived impact and potential to change behaviours and attitudes of men. Through a facilitated feedback session, men highlighted specific elements, phrases and opportunities to visualize the benefits of U=U they felt were especially compelling or memorable.

### Prototyping

The message transcriptions were compiled to develop a script that was around 60 seconds in duration (Box 1). In a second workshop, the outcomes from workshop one were presented to around 20 more men who were invited to rate and suggest improvements to the acceptability of the prototype message. Following the second HCD workshop, the engagement design and messaging was fine-tuned with technical experts from the Desmond Tutu HIV Foundation (DTHF) and the final message was summarised on printed invitation cards (Figure 3) and delivered verbally in a cluster randomized trial (18). The script was tested among n=7 men from the KMP district, as well as with n=4 HIV counsellors from the mobile clinic following a brief 10 question survey to assess comprehension and opinions about the messages as well as suggestions for improving the message.

#### Box 1

**Prototyped U=U testing invitation script**

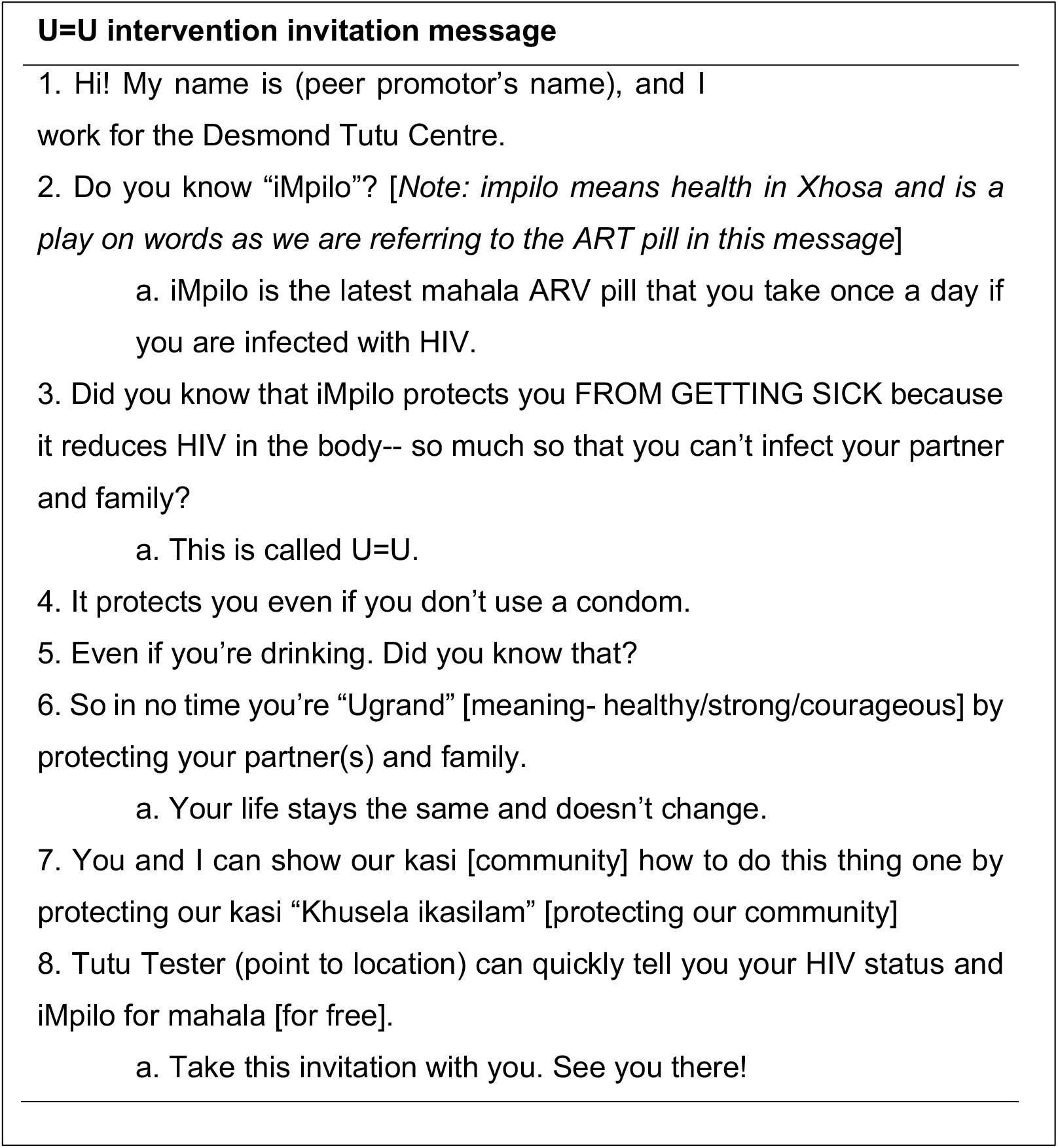

**Figure 3:**
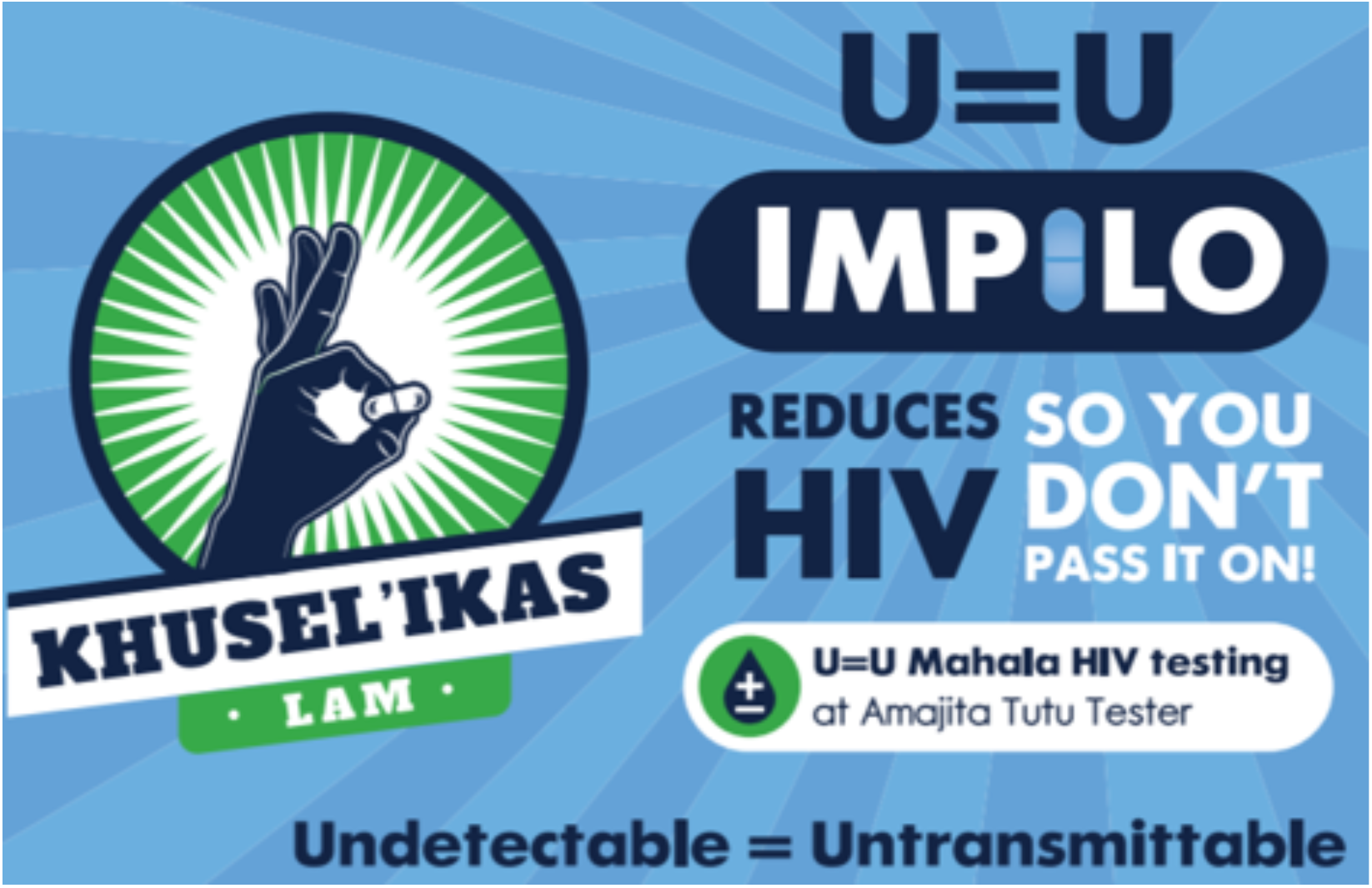
**Printed invitation to U=U intervention group with messaging around “Impilo (ART) reducing HIV so you don’t pass it on”**

## Results

The workshop enrolled n=39 adult men. Since ART was generally framed around treating sickness, men offered that messaging should be targeted towards desirable outcomes. The men expressed ideas around raising awareness, stating the benefits of ART, reducing fear, and ensuring men knew that the pill was easy to use. Participants emphasized three main insights in developing the U=U message; 1) introduce the modern antiretroviral pill, 2) positively redefine the man for whom the pill is intended, and 3) make the message simple (Box 2).

### Box 2

**Quotations from participant “pitches” during workshops**

*“When you are taking this tablet you cannot spread the virus to the other person*.*”*

*“This medicine boosts your body and the immune system*.*”*

*“Sir I wanted to tell you about something, there are new HIV pills which have been introduced they are very good and it has been proven 100%*.*”*

*“I explained to the potential customer (person) that I am having HIV positive and this pill helped me, by the way if you do not want to use a condom you cannot use it when you use this pill*.*”*

*“Get the HIV virus killer, so you don’t die. Finish and done!”*

*“The pill keeps killing so much virus in me so I can’t infect you baby*.*”*

*“The medication always kills so much virus in me. So, I will not infect you. No stress no worries, no pain and stress and no miseries*.*”*

*(rapping) “Umtholampilo (the pill) keeps on killing the virus in my body, so you can also take this Umtholampilo (the pill) so that you do not get infected*.*”*

*“That is me, I won’t be able to infect you with the disease*.*”*

*“It reduces the viral load in your body, that if only you have been tested positive, I am not saying that you are. When you are taking this tablet, you cannot spread the virus to the other person*.*”*

*“Most of the time this tablet is perfect for your health, but I would recommend that when you have time get tested so that you know your HIV status. Thank you very much for your time*.*”*

### Insights

#### Reintroduce the modern ART pill

Participants wanted to reintroduce the pill to ensure that men understand the benefits of taking ART. Specifically, participants wanted to convey that the pill ensured longevity. One of the participants stated, “The more you take the pill consistently, the longer you will live.” A participant who was not familiar with ART as a once-a-day pill said that when he became aware of the simplified regimen, “… it was music to my ears.”

#### Positively redefine the man for whom the pill is intended

Men spoke about how the U=U message would transform their identity. Since an HIV diagnosis was associated with worry about disclosure and deteriorating health, men stated that the U=U message should address those concerns. The participants said that the message could change the way that men saw themselves by reducing worry about a range of issues connected with concealing one’s HIV positive status;

> “You are able to disclose your status now because at least you won’t be infectious.”
>
> “No more fear, no more humiliation, no more stigma, no more loneliness, no more failure.”

Moreover, men spoke about the opportunity to reengage with aspirations, such as family life, having children and rearing a family, and social interactions. Having these values and aspirations were tentative in the aftermath of an HIV positive diagnosis. Participants stated that the U=U message could communicate that these aspirations could be normalised, stating;

> “You can grow your family without the fear of HIV.”
>
> “You are able to disclose your status now because at least you won’t be infectious.”

It was a relief for men to know that treatment had been simplified into a once-a-day regimen that reduced HIV viral load and prevented onward transmission. An even greater relief was the reduced social impact, including the option to drink alcohol and the assurance that an undetectable VL eliminated transmission – both of which reduced anxiety around these behaviours that were central to men’s identity and enjoyment.

#### Make the message simple

Men wanted a simple message to communicate the benefits of U=U. Men offered that the message should emphasize that 1) “you cannot spread the virus (HIV) to the other person” 2) and “(the pill) keeps on killing the virus.” In the messaging, participants also highlighted being able to drink alcohol, reduced side effects, and the once-a-day regimen that was simpler than previous regimens. Additionally, men wanted other men to know that the pill was free.

> “You can drink it with alcohol.”
>
> “The new ARV pill is taken anytime a day and it does not have side-effects.”
>
> “It won’t cost you a thing. It will sustain you forever.”

### Pitches

The men used the insights that they had generated to create a pitch of the U=U message. They wrote down their messages and delivered it to their peers. Examples of pitches are transcribed below:

> *“It’s a new tablet that has just come to the market, it is very helpful. Okay, I am also using this pill, it’s new on the market. Otherwise, I am also HIV positive brother, when I heard about this pill, I opted to use it. I have used it the way it was prescribed to me. It has raised me from the dead. So, can I explain it to you? Thank you for giving me your time brother. This pill is very helpful because I was bed bound and clueless, and then another guy came to explain to me about this tablet. I tried it while I was bed bound, so it helped me to get up. If you are interested on this tablet, I would like you to take it and try it on yourself and see how it works*.*”*
>
> *“Most of the time this tablet is perfect for your health, but I would recommend that when you have time get tested so that you know your HIV status. Thank you very much for your time*.*”*
>
> *“Umtholampilo (the pill) keeps on killing the virus in my body, so you can also take this Umtholampilo (the pill) so that you do not get infected*.*”*

### Intervention prototype

Insights and pitches were iteratively developed into a script and presentation that could be used in a brief face-to-face encounter to invite men to test. Early versions of the intervention incorporated rich content from workshops and an innovative visual presentation of individual and community VL using coloured glass beads. Both the length of the script and the use of the beads and other props were determined to be infeasible for purposes of pilot randomized study. Additional iterations, including field testing in the community, refined the script to 45-60 seconds and eliminated the glass bead presentation. The finalized prototype intervention was tested in a cluster randomized trial at five community testing sites with promising results (18).

## Discussion

Through a series of design workshops, we used participatory prototyping and human-centered design to translate the U=U (undetectable equal untransmittable) concept into a brief, upbeat, accurate invitation to seek HIV testing. In the workshop, men wanted to shift the emphasis of ART to highlight the beneficial outcomes of taking treatment. ART is often associated with complex regimens, side effects, and the associated hassle of visiting a clinic (19). To improve uptake and adherence, the workshop participants designed messages focusing on the desirable aspects of taking ART. Integral to the desirability of ART was making men aware that daily treatment was easier than it had been historically, especially that treatment did not impact on social behaviour, such as drinking alcohol. The participants also wanted to communicate that daily ART reduced HIV, which translated into being healthy again, with the benefit of longevity.

Stigma and denial remain significant problems in HIV endemic communities in South Africa. People (especially men) are disinclined to screen for illnesses such as HIV that are associated with severe illness or death. A common psychological barrier to testing was echoed in our workshops: men don’t want to find out if they have a dreaded disease like HIV because they think they will worry themselves to death. This is a particular challenging barrier given that many men become HIV-positive when they are young and feel healthy. Tailored messaging aimed at supporting desirable outcomes may circumvent the stigma and denial associated with testing and initiating ART.

Mental models about the meaning of an HIV diagnosis and the ability of ARVs to promote long and healthy survival have not been updated for the realities of modern ARVs. While the U=U messages we developed emphasize these new mental models, this has not been the case for public health campaigns South Africa to date. The men in our workshops who were not already on ART had the impression that ART still required many pills per day, had severe and numerous side effects, and offered only moderate treatment success.

### Study limitations

We sampled participants in from high disease burden communities in Cape Town. It is not known whether insights and pitches generated in this co-design process will generalize to men in different communities in South Africa or in other countries. Our study generated prototype designs for U=U informed interventions but require further field testing to assess effectiveness.

## Conclusion

Reversing and eradicating the HIV epidemic is dependent upon a treatment cascade that identifies and tests at-risk populations, ART start, adherence to treatment and consequent undetectable viral loads. Our design process to translate the U=U concept into testing invitations suggests that the benefits of ART have not yet been framed to MLHIV as a marketer might frame the benefits of a consumer product. U=U messaging that manages to capture this benefit and motivated men to seek HIV testing could be used as part of a multi-pronged intervention package to improve men’s entry into the prevention and treatment cascade. Considering multiple level interventions is imperative because they have been found to significantly increase rates of earlier testing, ART start, and drug adherence when compared to single mode interventions. Tailored messaging, mobile clinics, and early ART start may be a valuable addition to primary healthcare as they reach people earlier on in their infection, decongest conventional healthcare facilities, encourage healthy behaviour, and facilitate retaining clients over time.

## Data Availability

The qusalitative data is available upon request.

